# Digital Voice-Based Biomarker for Monitoring Respiratory Quality of Life: Findings from the Colive Voice Study

**DOI:** 10.1101/2023.11.11.23298300

**Authors:** Vladimir Despotovic, Abir Elbéji, Kevser Fünfgeld, Mégane Pizzimenti, Hanin Ayadi, Petr V. Nazarov, Guy Fagherazzi

**Affiliations:** Bioinformatics Platform, Department of Medical Informatics, Luxembourg Institute of Health, Strassen, Luxembourg; Deep Digital Phenotyping Research Unit, Department of Precision Health, Luxembourg Institute of Health, Strassen, Luxembourg; Multi-Omics Data Science, Department of Cancer Research, Luxembourg Institute of Health, Strassen, Luxembourg

**Keywords:** voice biomarker, respiratory quality of life, audio processing, deep learning

## Abstract

Regular monitoring of respiratory quality of life (RQoL) is essential in respiratory healthcare, facilitating prompt diagnosis and tailored treatment for chronic respiratory diseases. Voice alterations resulting from respiratory conditions create unique audio signatures that can potentially be utilized for disease screening or monitoring. Analyzing data from 1908 participants from the Colive Voice study, which collects standardized voice recordings alongside comprehensive demographic, epidemiological, and patient-reported outcome data, we evaluated various strategies to estimate RQoL from voice, including handcrafted acoustic features, standard acoustic feature sets, and advanced deep audio embeddings derived from pretrained convolutional neural networks. We compared models using clinical features alone, voice features alone, and a combination of both. The multimodal model combining clinical and voice features demonstrated the best performance, achieving an accuracy of 70.34% and an area under the receiver operating characteristic curve (AUROC) of 0.77; an improvement of 5% in terms of accuracy and 7% in terms of AUROC compared to model utilizing voice features alone. Incorporating vocal biomarkers significantly enhanced the predictive capacity of clinical variables across all acoustic feature types, with a net classification improvement (NRI) of up to 0.19. Our digital voice-based biomarker is capable of accurately predicting RQoL, either as an alternative to or in conjunction with clinical measures, and could be used to facilitate rapid screening and remote monitoring of respiratory health status.

## I. Introduction

MONITORING chronic respiratory diseases or other conditions that affect breathing is a foundation of respiratory healthcare. Telemonitoring solutions can help in reducing the workload of clinicians, decrease hospital admissions and shorten clinician response time, thus enabling more timely intervention. Remote monitoring is of utmost importance for identifying clinically relevant deterioration in the Respiratory Quality of Life (RQoL) and may be used as a prognostic tool for chronic respiratory conditions, such as Chronic Obstructive Pulmonary Disease (COPD) or asthma. A recent study proves that a decrease in RQoL by 4 points over a period of one year, measured by the St George’s Respiratory Questionnaire (SGRQ) [1], was associated with increased hospitalization and mortality. Besides SGRQ, other questionnaires have been also developed for estimating RQoL, including Chronic Respiratory Disease Questionnaire (CRDQ) [2], Breathing Problems Questionnaire (BPQ) [3], and VQ11 [4], just to name a few. Although questionnaires are considered essential in epidemiological studies, they are subjective, prone to biases and time-consuming; therefore, investigating alternative methods, such as analyzing voice characteristics, may provide valuable, scalable, easy-to-use solutions into assessing RQoL, requiring no invasive or cumbersome equipment, only a smartphone to record the voice.

The voice is a result of the airstream initiated in the lungs and respiratory airways, and passed through the larynx, causing the vibration of vocal folds, and furthermore through the oral and nasal cavity, where the sound is shaped and articulated. Respiratory diseases can alter the voice production process, resulting in distinctive changes in voice. Previous studies have shown that inspiratory closure of vocal folds, which causes refractory breathlessness, occurs frequently in COPD [5]. Changes in breathing and voice are highly correlated with altered lung function in patients with COPD [6], most likely affected by respiratory and muscle damage [7]. Acoustic features extracted from the speech are clearly distinctive during COPD exacerbation and stable periods [8], and are even distinguishable up to 7 days before the onset of symptoms [6]. Therefore, they could be used as an early warning system for COPD exacerbation.

Decreased voice-related quality of life, persistent cough and laryngeal dysfunction are also associated with up to 88% of patients with severe asthma [9]. Abnormal movements of vocal folds are caused by muscle tension in the vocal folds and larynx [9]. Vocal signatures extracted from voice recordings can be used to identify asthma worsening as a substitute to measures of lung function [10].

There are multiple advantages of monitoring respiratory diseases using voice recordings. The technology is non-invasive, cost-efficient and practical, requiring only smartphones to capture the voice; thus, could be used from patients’ homes for real-life remote monitoring in-between clinical visits or as a screening tool. Vocal biomarkers extracted from smartphone voice recordings were already used to identify pulmonary hypertension [11], and to monitor the recovery process of patients with influenza [12]. A number of studies for screening of COVID-19 from voice and cough smartphone recordings has recently appeared, either for the detection of COVID-19 [13], [14], [15], [16], [17], or for discriminating between the symptomatic and asymptomatic cases [18].

Contrary to the previous research works which were mostly focused on the identification and/or monitoring of respiratory diseases from voice, in this paper we investigated whether RQoL can be assessed from voice features. Instead of targeting a single respiratory disease, we analyze RQoL in a general population containing participants with multiple respiratory conditions (e.g. asthma, COPD) as well as participants with no history of respiratory diseases, by stratifying them according to VQ11 scores, and comparing voice signatures extracted from sustained vowel phonation recordings. As an objective measure, vocal biomarkers can increase the reliability of screening based only on subjective self-reports. To support this hypothesis, we used data from the international, multilingual Colive Voice initiative to show that voice can be utilized as a universal biomarker for monitoring chronic respiratory conditions, either alone, or in addition to clinical parameters extracted from self-administered questionnaires. To our knowledge, this is the first study that proposes a multimodal approach combining voice features with clinical data.

## II. Material and methods

### A. Study design

Colive Voice^1^ is an international digital health study established and led by the Luxembourg Institute of Health which aims at identifying vocal biomarkers for remote monitoring and screening of various chronic diseases and frequent health symptoms. The multilingual audio databank is collected in four languages (English, French, German and Spanish) and contains recordings of multiple vocal tasks, including sustained vowel phonation, coughing, breathing, reading and counting. Voice recordings are associated with annotated clinical and demographic data, providing an in-depth patient characterization with validated disease-specific questionnaires on symptoms, treatments and quality of life. Colive Voice has been hosted online since June 2021 and is open for participation to anyone, under the condition that: 1) they sign the consent form and 2) they are at least 15 years old.

The study has been approved by the National Research Ethics Committee in Luxembourg (N° 202103/01) in March 2021. Informed written consent was obtained electronically via the Colive Voice application from all participants in the study. The Colive Voice study protocol is also registered on ClinicalTrials.gov (NCT04848623).

Part of the study is dedicated to investigation of RQoL in the general population from voice recordings, accompanied with annotations of RQoL via self-administered VQ11 questionnaire, as well as clinical and demographic data.

Unlike SGRQ and BPQ, which are extensive (76 items in SGRQ and 33 items in BPQ) with complex scoring, making them unsuitable for repeated evaluations in clinical practice as well as a regular use in real life, VQ11 is a brief questionnaire with only 11 items distributed across functional omponents (3 items), psychological components (4 items) and social components (4 items). Although much simpler and faster to record, VQ11 shows high correlation with SGRQ [4]. Each item in VQ11 is represented by five categories (not at all, a little, moderately, much, extremely) which reflect the participant’s feeling about the statement associated with a particular item, and can be represented by a value from 1 to 5. The total score is obtained by summing all individual items, leading to a score between 11 and 55 with lower value indicating better RQoL [4]. We stratify the participants in the study into two categories using the cut-off VQ11 score of 22: 1) Impaired RQoL (*V Q*11 ≥ 22), and 2) Normal RQoL (*V Q*11 *<* 22) [19], [20].

Since the number of participants with impaired RQoL was significantly lower than the normal RQoL, we generate a balanced dataset matched by age and gender composed of 1908 sustained vowel recordings in total, equally distributed between two groups.

A full workflow of RQoL monitoring from data acquisition to the prediction of RQoL is shown in Figure 1.

**Fig. 1:**
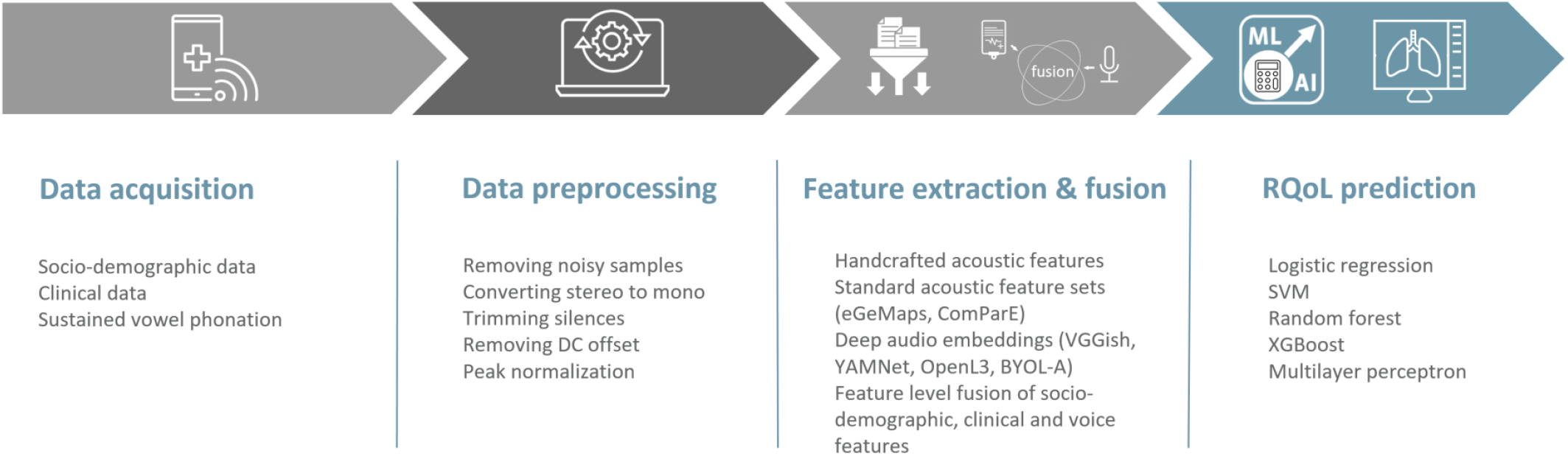
Workflow of RQoL monitoring

### B. Data acquisition and preprocessing

Participants were recruited via an online crowdsourced campaign or through partnerships with various patient associations, academic institutions, hospitals, or other research initiatives (including Les Sentinelles and the ComPaRe study, AP-HP). The full list of partners is available on the Colive Voice website. Participants were invited to use an app^2^ accessible from participants’ devices equipped with microphones (smartphone, tablet or laptop). Collected information is composed of socio-demographic and clinical data acquired via participants’ self-reported questionnaires and voice recordings.

Socio-demographic data contains information about body mass index (BMI) and smoking habits, while clinical data contains information about day and night coughing, chest pain, sore throat, as well as associated diseases such as asthma and COPD. Categorical variables were encoded as one-hot representations, leading to 23 features in total.

Voice recordings are acquired in the form of sustained vowel phonation (/a/ vowel) produced at a comfortable pitch and loudness as long as possible. Vowel phonation is selected since it provides valuable information about the pulmonary function, and in addition, it is less susceptible to language bias, which may be present in the multi-lingual data collection. Reduced pulmonary function leads to decreased airflow necessary to support phonation [21], which in turn reflects in reduced RQoL.

Participants were advised to make voice recordings in a quiet environment without the external noise in order to preserve high-quality recordings. However, given that data is collected in uncontrolled conditions and to account for the challenges related to the use of different devices, microphones, and recording conditions for data collection, audio preprocessing using a proprietary pipeline was performed to harmonize the recordings and prepare them for the subsequent steps.

### C. Statistical analysis

We utilized an independent two-tailed t-test to compare the means of groups with normal and impaired RQoL for continuous variables. For categorical variables, a chi-square test was used. A p-value less than 0.05 indicates a statistically significant difference. Only variables that were statistically significant were used as socio-demographic and clinical features in further processing. Table I provides a summary of the study population characteristics.

**TABLE I:** Study population characteristics.

| | | Total | | | Normal RQoL ($VQ_{11} < 22$) | | | Impaired RQoL ($VQ_{11} \geq 22$) | | | p value |
| --- | --- | --- | --- | --- | --- | --- | --- | --- | --- | --- | --- |
| Participants |  | 1908 |  |  | 954 (50%) |  |  | 954 (50%) |  |  | NA |
| Mean VQ11 score |  | 21.6 (8.2) |  |  | 15 (3) |  |  | 28.3 (6.1) |  |  | < 0.0001 |
| Gender |  | F | M | O | F | M | O | F | M | O | 1 |
|  |  | 1280<br>(67.1%) | 608<br>(31.9%) | 20<br>(1%) | 640<br>(67.1%) | 304<br>(31.9%) | 10<br>(1%) | 640<br>(67.1%) | 304<br>(31.9%) | 10<br>(1%) |  |
| Age |  | 42.4 (14.2) |  |  | 42.4 (14.1) |  |  | 42.5 (14.2) |  |  | 0.948 |
| BMI [kg/m<sup>2</sup>] | Underweight | 66 (3.5%) |  |  | 35 (3.7%) |  |  | 31 (3.2%) |  |  | < 0.0001 |
|  | Normal weight | 792 (41.5%) |  |  | 490 (51.3%) |  |  | 302 (31.7%) |  |  |  |
|  | Overweight | 466 (24.4%) |  |  | 224 (23.5%) |  |  | 242 (25.4%) |  |  |  |
|  | Obesity | 601 (30.6%) |  |  | 205 (21.5%) |  |  | 379 (39.7%) |  |  |  |
| Smoking status | Not at all | 1533 (80.4%) |  |  | 806 (84.5%) |  |  | 727 (76.2%) |  |  | < 0.0001 |
|  | Less than daily | 98 (5.1%) |  |  | 50 (5.2%) |  |  | 48 (5%) |  |  |  |
|  | Daily | 277 (14.5%) |  |  | 98 (10.3%) |  |  | 179 (18.8%) |  |  |  |
| Day coughing | No | 1181 (61.9%) |  |  | 704 (73.8%) |  |  | 477 (50%) |  |  | < 0.0001 |
|  | Transient | 597 (31.3%) |  |  | 235 (24.6%) |  |  | 362 (38%) |  |  |  |
|  | Frequent | 130 (6.8%) |  |  | 15 (1.6%) |  |  | 115 (12%) |  |  |  |
| Night coughing | No | 1414 (74.1%) |  |  | 802 (84%) |  |  | 612 (64.2%) |  |  | < 0.0001 |
|  | Transient | 396 (20.8%) |  |  | 137 (14.4%) |  |  | 259 (27.1%) |  |  |  |
|  | Frequent | 98 (5.1%) |  |  | 15 (1.6%) |  |  | 83 (8.7%) |  |  |  |
| Chestpain | Yes | 191 (10%) |  |  | 43 (4.5%) |  |  | 148 (15.5%) |  |  | < 0.0001 |
| Sore throat | Yes | 190 (10%) |  |  | 71 (7.4%) |  |  | 119 (12.5%) |  |  | 0.0002 |
| Asthma | Yes | 306 (16%) |  |  | 118 (12.4%) |  |  | 188 (19.7%) |  |  | < 0.0001 |
| COPD | Yes | 73 (3.8%) |  |  | 18 (1.9%) |  |  | 55 (5.8%) |  |  | < 0.0001 |

F - Female; M - Male; O - Other; NA - Not Applicable; BMI - Body Mass Index; COPD - Chronic Obstructive Pulmonary Disease; $p < 0.05$ is considered statistically significant.

### D. Feature extraction and fusion

We first extracted a set of 72 handcrafted audio features, that contain time domain, spectral, cepstral, prosodic, and nonlinear dynamics features (Table II). Audio features were extracted using Surfboard [22], a Python library for feature extraction with application to the medical domain, as well as Parselmouth [23], a Python interface to Praat. We selected the audio features that are shown to be relevant for vocal biomarker research across multiple diseases.

**TABLE II:** Handcrafted audio features.

| ID | Feature | Domain | Computation parameters |
| --- | --- | --- | --- |
| 1-26 | MFCC | Cepstral | Mean and standard deviation of 13 MFCCs |
| 27 | RMS power | Time | None |
| 28 | Zero crossing rate | Time | None |
| 29 | Crest factor | Time | None |
| 30 | Dominant frequency | Spectral | None |
| 31 | Spectral centroid | Spectral | None |
| 32 | Spectral rolloff | Spectral | None |
| 33 | Spectral spread | Spectral | None |
| 34 | Spectral skewness | Spectral | None |
| 35 | Spectral kurtosis | Spectral | None |
| 36 | Spectral bandwidth | Spectral | None |
| 37 | Spectral flatness | Spectral | None |
| 38 | Spectral standard deviation | Spectral | None |
| 39 | Spectral slope | Spectral | None |
| 40 | Spectral decrease | Spectral | None |
| 41 | Maximum phonation time | Time | None |
| 42-44 | Aperiodicity features | Time | Fraction of locally unvoiced frames, Number of voice breaks, Degree of voice breaks |
| 45-54 | Tremor | Time | Frequency contour magnitude, amplitude contour magnitude, frequency tremor cyclicity, amplitude tremor cyclicity, frequency tremor frequency, amplitude tremor frequency, frequency tremor intensity index, amplitude tremor intensity index, frequency tremor power index, amplitude tremor power index |
| 55-59 | Jitter | Time | Local, local absolute, RAP, ppq5, ddp |
| 60-65 | Shimmer | Time | Local, local [dB], apq3, apq5, apq11, dda |
| 66 | Detrended fluctuation analysis | Nonlinear dynamics | None |
| 67 | Shannon entropy | Nonlinear dynamics | None |
| 68 | Harmonics to noise ratio | Time | None |
| 69-70 | Fundamental frequency (F0) | Prosodic | Mean, standard deviation |
| 71-72 | F0 contour | Prosodic | Mean, standard deviation |

MFCC - Mel-frequency cepstral coefficients; RMS - Root mean square; RAP - Relative average perturbation; ppq - Period perturbation quotient; ddp - Difference of differences of periods; apq - Amplitude perturbation quotient; dda - Difference of differences of amplitudes.

In addition to this, we used standard audio feature sets, i.e. extended Geneva Minimalistic Acoustic Parameter Set (eGeMAPS) [24] and ComParE, extracted using the openSMILE [25]. The eGeMAPS is a minimalistic set of acoustic parameters for paralinguistic or clinical speech analysis which is composed of 88 energy/amplitude, frequency, spectral and temporal features, as well as statistical functionals applied to them (arithmetic mean, standard deviation, percentile). ComParE is a brute force audio feature set that contains 65 low-level acoustic descriptors and various statistical functionals applied to them, leading to a total of 6737 audio features.

Finally, we experiment with 4 different types of deep audio embeddings, i.e. VGGish [26], YAMNet, OpenL3 [27], and BYOL-A [28], which are state-of-the-art general audio features pretrained on large audio collections that are successfully used for a number of downstream tasks. Characteristics of different audio embeddings are provided in Table III.

**TABLE III:**
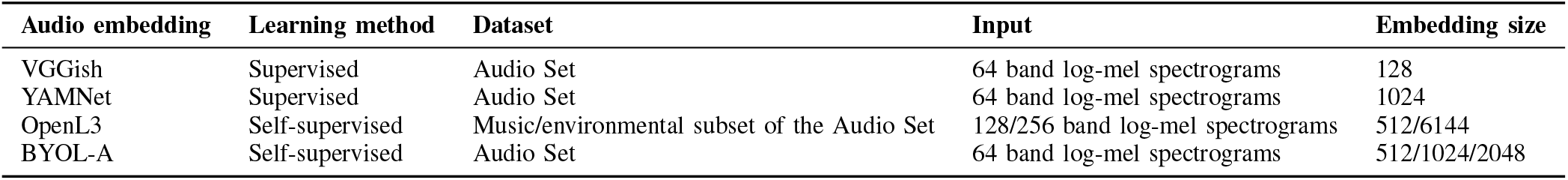
Characteristics of the deep audio embeddings.

| Audio embedding | Learning method | Dataset | Input | Embedding size |
| --- | --- | --- | --- | --- |
| VGGish | Supervised | Audio Set | 64 band log-mel spectrograms | 128 |
| YAMNet | Supervised | Audio Set | 64 band log-mel spectrograms | 1024 |
| OpenL3 | Self-supervised | Music/environmental subset of the Audio Set | 128/256 band log-mel spectrograms | 512/6144 |
| BYOL-A | Self-supervised | Audio Set | 64 band log-mel spectrograms | 512/1024/2048 |

VGGish is a pretrained convolutional neural network (CNN) mostly inspired by the VGG network used in computer vision. The network is adapted to accept 96×64 bin log-mel spectrograms at its input and extracts 128-dimensional embeddings from 960 ms segments of an audio signal. YAMNet employs the Mobilenet v1 depthwise separable convolution architecture used with the same input as VGGish, but outputs 1024-dimensional embeddings for each 960 ms audio segment. Both VGGish and YAMNet are pretrained on the large-scale Audio Set dataset for audio event classification which contains more than 2 million of 10 s YouTube clips of sounds classified into 632 audio events. To summarize features across different audio segments and output the equal size feature vectors from recordings of different lengths average pooling was used.

OpenL3 uses CNN-based L3-Net for self-supervised learning via audio-visual correspondence, to learn whether a particular video frame corresponds to an audio frame; thus, requiring no annotations. The model is pretrained on two subsets of Audio Set, i.e. music and environmental subset, containing 296K and 195K clips respectively, and uses either 128 or 256 band Mel-spectrograms at the input, while the output audio embedding is 512 or 6144-dimensional vector for each 1s audio segment. We use a model pretrained on an environmental subset, with 256 band Mel-spectrograms and 6144-dimensional embeddings.

BYOL-A uses the Bootstrap Your Own Latent (BYOL) method for self-supervised learning of general-purpose image representations, adapted to work with audio. Normalized 96×64 bin log-mel spectrograms are used as an input, and two augmented versions of the input are created by shifting pitch and stretching time, which are further fed into two parallel networks (online and target network). The online network predicts the output representation of the target network, which is then iteratively updated as the exponential moving average of the parameters of the online network. The model is pretrained on the Audio Set dataset and produces 512, 1024 or 2048-dimensional general-purpose audio embeddings. We use 2048-dimensional embeddings.

In addition to voice features, we extracted the demographic/clinical data relevant for RQoL from the subjective self-reports. We used socio-demographic variables that were found statistically significant (BMI, smoking habits), symptoms (day and night coughing, chest pain, sore throat), and associated diseases that can affect RQoL (asthma, COPD), as shown in Table I. All categorical variables were encoded as one-hot representations, except for ensemble-based models (Random Forest, Extreme Gradient Boosting), where single feature representation was kept. One-hot encodings produce sparse feature vectors, which are not suitable for tree-based models, since splitting on such features produces a small gain, and is typically ignored in favor of continuous variables. Features were standardized before feeding them to classification models to put them on the same scale, i.e. all features have zero mean and unit standard deviation.

Given that the size of the audio feature vectors is substantially larger than the size of socio-demographic/clinical features (up to 250 times larger for ComParE features), Principal Component Analysis (PCA) was applied to audio embeddings prior to data fusion, to reduce their dimensionality to the first 23 principal components that explain most of the variance, and to put the features from different modalities to equal dimension.

### E. RQoL prediction

Features extracted in the previous section were fed into several classifiers: Logistic Regression (LR), Support Vector Machines (SVM), Random Forest (RF), Extreme Gradient Boosting (XGBoost), and Multilayer Perceptron (MLP).

LR with L2 regularization is used to handle overfitting, as well as SVM with radial basis function kernel, where the model hyperparameters, i.e. the regularization parameter *C* and the kernel coefficient *γ* are optimized using a grid search. Two ensemble models include RF and XGBoost. RF was composed of 500 fully grown trees (optimal number of trees was determined after hyperparameter tuning), expanded until all leaves were pure or contained less than 2 samples, with Gini index as the criterion for splitting the node, and the number of features at each split equal to the square root of the total number of features. All models are implemented using the scikit-learn 1.1.3 Python library.

XGBoost is a flexible and distributed gradient boosting algorithm, that allows for custom loss functions, as well as regularization techniques to mitigate the overfitting. We use XGBoost with 500 trees, L2 regularization and log loss objective function. XGBoost is implemented using the xgboost 1.5.0 Python library.

MLP was composed of two hidden layers with 256 neurons each and a ReLU activation function, followed by dropout layers for preventing overfitting with a dropout rate equal to 0.3, and an output layer with a sigmoid activation function is utilized in this paper. We used Adam optimizer, binary cross entropy loss function, batch size equal to 32, while the optimal learning rate (0.0001) and the number of epochs (30) are determined via grid search. Note that Adam has an adaptive per-parameter learning rate, which is computed using the initial learning rate as an upper limit. MLP is implemented using Tensorflow 2.9.1.

### F. Evaluation

For evaluation of the model performance we use accuracy, sensitivity, specificity, area under the receiver operating characteristic curve (AUROC), Brier score and net reclassification index (NRI).

Accuracy is the ratio of the number of correctly classified observations and the total number of observations. Sensitivity (true positive rate, recall) is the proportion of participants detected with impaired RQoL (true positives) among those who have impaired RQoL (true positives + false negatives), and shows the model’s ability to correctly identify cases. Specificity (true negative rate) is the proportion of participants detected with normal RQoL (true negatives) among those who have normal RQoL (true negatives + false positives), and refers to the model’s ability to correctly identify healthy controls. ROC curve plots sensitivity against false negative rate (1-specificity) at different classification thresholds, while AUROC is an aggregated performance measure which summarizes ROC curve, with a value of 0.5 denoting random guess, and 1 denoting perfect classification.

To assess model calibration, i.e. the consistency between the predicted probability and the observations, Brier score was used, which is the mean squared deviation of the predicted probability from the actual target. It is a value between 0 and 1, with a lower value indicating a better model.

Given the size of the dataset, to get the reliable and robust performance estimates and preserve the class distribution across folds, we used stratified 5-fold cross-validation [29]. Data is split into five folds, four merged and used for training, and the remaining one for testing. The process is repeated 5 times, so that each fold was used exactly once for testing, and the performance is then averaged over all folds.

Finally, since our aim was to quantify how much voice-related information can improve the reliability of RQoL screening on top of standard clinical features, we used NRI to estimate the improvement in performance due to adding vocal biomarkers to a set of socio-demographic and clinical predictors. The value can range from -2 to 2, with bigger value indicating larger improvement.

## III. Results

### A. Evaluation of RQoL from socio-demographic/clinical data

Before evaluating the relevance of vocal biomarkers for estimating RQoL, we set up a baseline experiment where only socio-demographic data (BMI, smoking habits) and clinical data (day and night coughing, chest pain, sore throat, as well as associated diseases such as asthma and COPD) from the participants’ self-reports were used for prediction of RQoL status. Categorical variables were encoded as one-hot representations, leading to 23 features in total. Performance is averaged over 5 folds (Table IV), with the best AUROC of 0.70, and accuracy of 64.1% obtained using LR classifier.

**TABLE IV:**
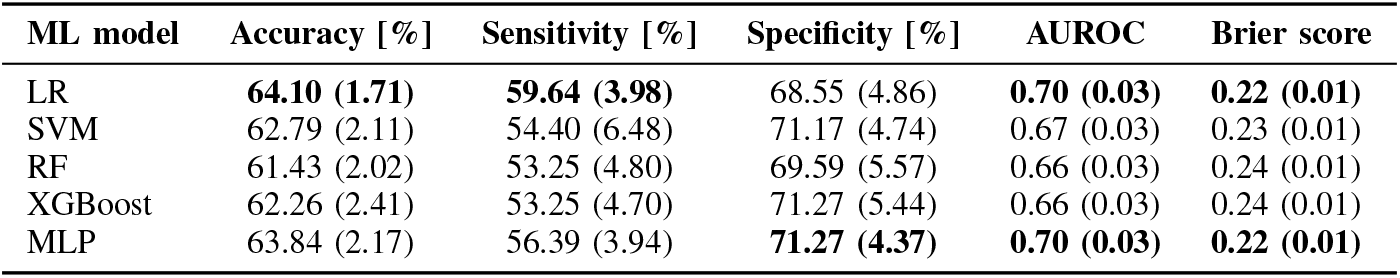
RQoL assessment based on socio-demographic and clinical features.

| ML model | Accuracy [%] | Sensitivity [%] | Specificity [%] | AUROC | Brier score |
| --- | --- | --- | --- | --- | --- |
| LR | **64.10 (1.71)** | **59.64 (3.98)** | 68.55 (4.86) | **0.70 (0.03)** | **0.22 (0.01)** |
| SVM | 62.79 (2.11) | 54.40 (6.48) | 71.17 (4.74) | 0.67 (0.03) | 0.23 (0.01) |
| RF | 61.43 (2.02) | 53.25 (4.80) | 69.59 (5.57) | 0.66 (0.03) | 0.24 (0.01) |
| XGBoost | 62.26 (2.41) | 53.25 (4.70) | 71.27 (5.44) | 0.66 (0.03) | 0.24 (0.01) |
| MLP | 63.84 (2.17) | 56.39 (3.94) | **71.27 (4.37)** | **0.70 (0.03)** | **0.22 (0.01)** |

We also presented the feature importance based on the mean impurity decrease for the RF model in Figure 2, revealing that BMI is the most important socio-demographic variable, followed by clinical symptoms related to day and night coughing.

**Fig. 2:**
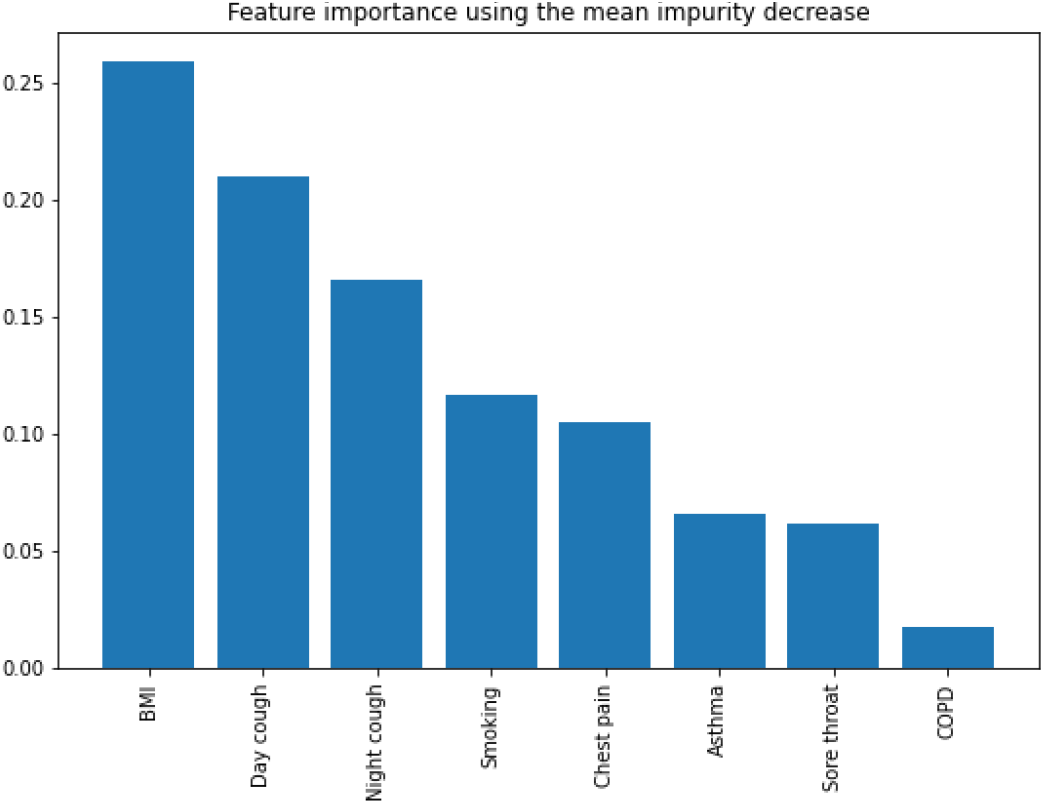
Feature importance for socio-demographic and clinical features based on the mean impurity decrease

### B. Evaluation of RQoL from voice recordings

We investigated whether voice related information could be used as a digital biomarker for RQoL. To that end, we extracted a set of handcrafted audio features (Table II), as well as two widely used general audio feature sets (eGeMAPS and ComParE). In addition to this, four state-of-the-art deep audio embeddings are evaluated (VGGish, YAMNet, OpenL3, BYOL-A) which proved to be highly competitive across multiple audio tasks. The features were either fed directly to the classifier, or in case of deep audio embeddings after applying Principal Component Analysis (PCA) to reduce the dimensionality of feature vectors. The results for assessment of RQoL from voice were provided in Table V, with the best performance reaching AUROC equal to 0.7 and accuracy of 65.57% using BYOL-A deep audio embeddings. BYOL-A substantially outperforms all other feature extraction techniques by over 2%.

**TABLE V:** RQoL assessment based on handcrafted voice features, standard acoustic feature sets, and deep audio embeddings.

| ML model | Features | Accuracy [%] | Sensitivity [%] | Specificity [%] | AUROC | Brier score |
| --- | --- | --- | --- | --- | --- | --- |
| LR | Handcrafted | 60.48 (3.42) | 56.30 (4.75) | 64.68 (4.64) | 0.67 (0.03) | 0.23 (0.01) |
|  | eGeMAPS | 59.54 (2.37) | 53.25 (2.08) | 65.82 (3.34) | 0.64 (0.03) | 0.23 (0.01) |
|  | ComParE | 62.58 (1.84) | 54.62 (3.17) | 70.55 (1.49) | 0.67 (0.02) | 0.23 (0) |
|  | VGGish | 61.69 (1.82) | 59.86 (3.51) | 63.52 (1.15) | 0.66 (0.02) | 0.23 (0.01) |
|  | YAMNet | 61.53 (2.63) | 53.67 (2.74) | 69.39 (3.32) | 0.67 (0.01) | 0.23 (0.01) |
|  | OpenL3 | 62.16 (2.86) | 56.71 (4.04) | 67.61 (2.25) | 0.67 (0.03) | 0.23 (0.01) |
|  | BYOL-A | **65.57 (1.66)** | **59.96 (3.46)** | 71.17 (1.92) | **0.70 (0.02)** | **0.22 (0)** |
| SVM | Handcrafted | 62.11 (3.84) | 55.15 (4.65) | 69.07 (4.56) | 0.67 (0.04) | 0.23 (0.01) |
|  | eGeMAPS | 58.91 (1.91) | 45.91 (2.97) | 71.90 (3.95) | 0.63 (0.02) | 0.23 (0) |
|  | ComParE | 63.37 (1.49) | 47.70 (3.58) | **79.04 (1.47)** | 0.66 (0.03) | 0.23 (0.01) |
|  | VGGish | 60.74 (1.38) | 52.62 (2.13) | 68.87 (1.79) | 0.66 (0.02) | 0.23 (0) |
|  | YAMNet | 62.06 (1.69) | 48.12 (2.55) | 76.00 (2.30) | 0.67 (0.01) | 0.23 (0) |
|  | OpenL3 | 63.58 (2.10) | 57.23 (2.46) | 69.92 (2.11) | 0.67 (0.03) | 0.23 (0.01) |
|  | BYOL-A | 63.99 (1.58) | 52.2 (3.10) | 75.79 (2.37) | 0.69 (0.02) | 0.22 (0.01) |
| RF | Handcrafted | 60.64 (2.95) | 58.49 (4.38) | 62.79 (4.17) | 0.65 (0.03) | 0.23 (0.01) |
|  | eGeMAPS | 58.65 (2.17) | 55.45 (2.96) | 61.85 (2.03) | 0.62 (0.02) | 0.24 (0) |
|  | ComParE | 61.16 (1.92) | 55.98 (3.06) | 66.35 (2.29) | 0.64 (0.03) | 0.23 (0.01) |
|  | VGGish | 60.38 (1.52) | 57.23 (1.77) | 63.52 (1.99) | 0.64 (0.02) | 0.23 (0) |
|  | YAMNet | 61.79 (1.45) | 57.34 (3.49) | 66.25 (2.11) | 0.66 (0.02) | 0.23 (0) |
|  | OpenL3 | 62.26 (1.66) | 55.77 (3.26) | 68.76 (0.75) | 0.66 (0.03) | 0.28 (0.01) |
|  | BYOL-A | 62.37 (2.50) | 58.18 (3.75) | 66.56 (1.89) | 0.67 (0.03) | 0.23 (0.01) |
| XGBoost | Handcrafted | 58.07 (2.75) | 57.23 (3.05) | 58.91 (4.98) | 0.62 (0.02) | 0.31 (0.01) |
|  | eGeMAPS | 57.91 (1.18) | 56.50 (1.32) | 59.33 (2.75) | 0.61 (0.01) | 0.32 (0.01) |
|  | ComParE | 58.12 (2.28) | 54.52 (4.90) | 61.74 (2.39) | 0.62 (0.03) | 0.32 (0.02) |
|  | VGGish | 56.87 (1.12) | 56.40 (1.49) | 57.34 (1.58) | 0.60 (0.02) | 0.33 (0.01) |
|  | YAMNet | 58.76 (2.98) | 57.97 (4.36) | 59.54 (3.17) | 0.63 (0.02) | 0.32 (0.02) |
|  | OpenL3 | 57.71 (2.70) | 54.72 (2.78) | 60.70 (4.32) | 0.63 (0.02) | 0.32 (0.01) |
|  | BYOL-A | 58.75 (2.72) | 56.40 (3.15) | 61.11 (3.86) | 0.63 (0.02) | 0.32 (0.01) |
| MLP | Handcrafted | 61.58 (3.10) | 58.08 (5.43) | 65.09 (3.01) | 0.66 (0.03) | 0.23 (0.01) |
|  | eGeMAPS | 60.95 (1.56) | 52.62 (1.50) | 69.29 (2.88) | 0.64 (0.02) | 0.24 (0.01) |
|  | ComParE | 58.81 (2.30) | 54.94 (7.62) | 62.68 (5.99) | 0.63 (0.03) | 0.25 (0.01) |
|  | VGGish | 60.22 (2.63) | 57.03 (5.72) | 63.42 (3.63) | 0.65 (0.03) | 0.23 (0.01) |
|  | YAMNet | 62.26 (0.55) | 54.72 (2.23) | 69.81 (3.00) | 0.67 (0.01) | 0.23 (0) |
|  | OpenL3 | 60.69 (1.72) | 56.92 (5.83) | 64.47 (2.83) | 0.64 (0.02) | 0.24 (0.01) |
|  | BYOL-A | 62.53 (3.27) | 54.20 (5.21) | 70.86 (5.05) | 0.67 (0.03) | 0.23 (0.01) |

To highlight the characteristics of sustained vowel phonation labeled with normal and impaired RQoL, we showed in Figure 3 spectrograms of two participants matched by age and gender (males, 67 years old): one with normal RQoL without the history of pulmonary diseases, but with a diagnosed COVID-19 more than 3 weeks before the recording was made; and one with extremely impaired RQoL (VQ11 score: 46) diagnosed with asthma-COPD overlap syndrome. Even though the normal RQoL example is actually a boundary case (VQ11 score: 21, cut-off value 22), the differences in spectrograms are clearly visible. While the normal RQoL recording is represented by uninterrupted phonation, with clearly distinctive harmonics (Figure 3a), impaired RQoL recording is characterized by strangled voice with multiple stoppages and voice breaks, and increased energy areas in higher frequency bands, which are most likely caused by aperiodic noise produced at a glottal constriction (Figure 3b). Furthermore, the absence of higher harmonics above 1kHz can be observed throughout the spectrogram, and as phonation progresses, even the adjacent lower harmonics become smeared and more difficult to distinguish. However, for the impaired RQoL voice recordings with VQ11 score closer to cut-off value, the differences are not so distinct.

**Fig. 3:**
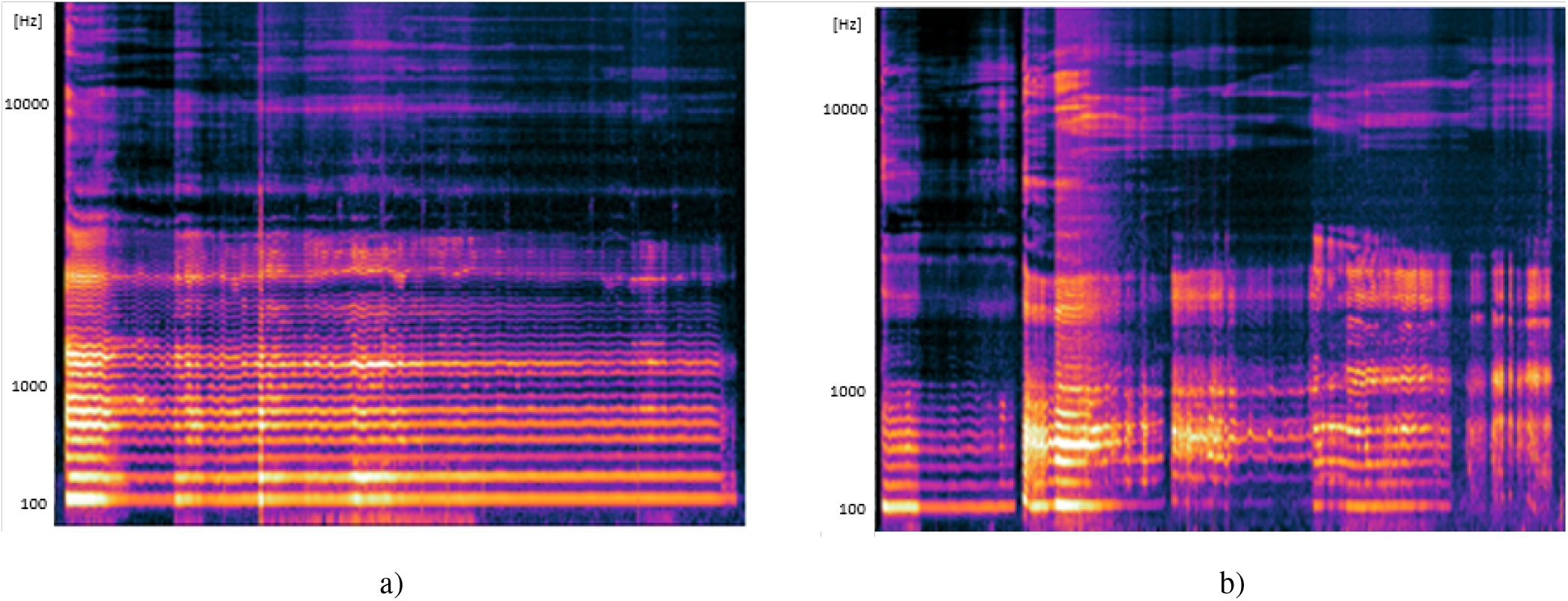
Spectrograms of sustained vowel phonation of participants matched by age and gender (male, age 67) with a) normal RQoL (VQ11 score: 21); and b) impaired RQoL (VQ11 score: 46). Normal RQoL spectrogram is represented by uninterrupted phonation, with clearly distinctive harmonics. Impaired RQoL spectrogram is characterized by strangled voice with multiple stoppages and voice breaks, and increased energy areas in higher frequency bands. The absence of higher harmonics above 1kHz can be observed, and as phonation progresses, the adjacent lower harmonics become smeared and more difficult to distinguish.

### C. Evaluation of RQoL from fused socio-demographic/clinical data and voice recordings

By fusing socio-demographic/clinical with voice features, we can quantify how much voice features can boost the performance of the socio-demographic and clinical data, uncovering the full potential of the multimodal data fusion. The results for the assessment of RQoL from multimodal features are provided in Table VI, whereas the comparison of the best-performing machine learning model for the socio-demographic/clinical features only, voice features only and multimodal features obtained after their fusion is presented in Figure 4. By using intermediate fusion (feature level fusion) we show that clinical data extracted from questionnaires and voice features obtained as the higher-level representations extracted from raw audio signals are complementary, leading to a substantial performance boost (accuracy equal to 70.34% and AUC equal to 0.77 using the combination of BYOL-A audio embeddings and socio-demographic/clinical features). Note that specificity is, in general, higher than sensitivity for all models, i.e. the models are still better at predicting normal than impaired RQoL. This is also visible from the confusion matrix of the best-performing model (fused BYOL-A and socio-demographic/clinical features, trained with LR classifier) shown in Figure 5a, where it is clear that the number of false negatives is substantially larger than the number of false positives. Using the Brier score as a measure of calibration, the same multimodal model achieves the lowest average Brier score over all folds equal to 0.19 with a nearly linear calibration curve, as shown in Figure 5b. Figure 5c displays the ROC curve of the best-performing model.

**Fig. 4:**
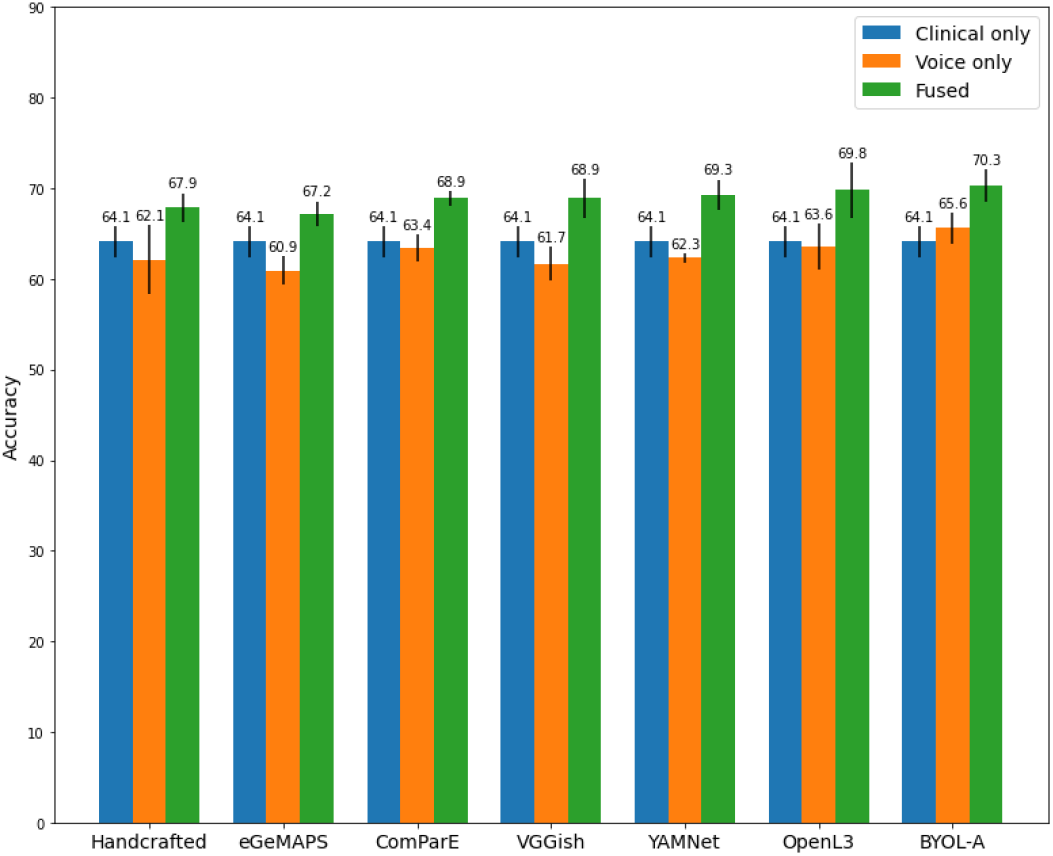
Accuracy with the best-performing machine learning model for socio-demographic/clinical features only, voice features only and fused clinical and voice (multimodal) features. Models with both clinical and voice data (“Fused”) systematically outperformed models where clinical variables only or voice features only were used. Error bars represent the standard deviation.

**TABLE VI:** RQoL assessment based on fused socio-demographic/clinical and voice features.

| ML model | Features | Accuracy [%] | Sensitivity [%] | Specificity [%] | AUROC | Brier score | NRI |
| --- | --- | --- | --- | --- | --- | --- | --- |
| LR | Handcrafted | 67.77 (2.07) | 63.42 (2.47) | 72.12 (2.06) | 0.74 (0.01) | 0.20 (0.01) | 0.15 |
|  | eGeMAPS | 67.14 (2.24) | 61.53 (2.98) | 72.74 (3.27) | 0.73 (0.02) | 0.21 (0.01) | 0.16 |
|  | ComParE | 68.92 (0.85) | 64.26 (2.17) | 73.59 (0.78) | 0.75 (0.02) | 0.20 (0.01) | 0.14 |
|  | VGGish | 68.92 (2.18) | 65.62 (3.55) | 72.22 (1.56) | 0.75 (0.02) | 0.20 (0.01) | 0.14 |
|  | YAMNet | 69.34 (1.65) | 64.05 (2.27) | 74.63 (2.60) | 0.76 (0.02) | 0.20 (0.01) | 0.17 |
|  | OpenL3 | 69.76 (2.82) | 65.83 (3.15) | 73.69 (2.53) | 0.76 (0.02) | 0.20 (0.01) | 0.16 |
|  | BYOL-A | **70.34** (1.82) | **66.78** (4.60) | 73.90 (1.44) | **0.77** (0.02) | **0.19** (0.01) | 0.10 |
| SVM | Handcrafted | 67.87 (1.24) | 60.38 (1.80) | 75.37 (2.40) | 0.74 (0.01) | 0.20 (0) | 0.12 |
|  | eGeMAPS | 67.24 (1.38) | 56.08 (1.75) | 78.41 (1.67) | 0.73 (0.01) | 0.21 (0.01) | **0.19** |
|  | ComParE | 66.98 (1.79) | 58.70 (2.49) | 75.26 (2.07) | 0.72 (0.03) | 0.21 (0.01) | 0.13 |
|  | VGGish | 68.66 (1.41) | 61.01 (2.57) | 76.31 (1.97) | 0.75 (0.02) | 0.20 (0.01) | 0.16 |
|  | YAMNet | 68.76 (1.94) | 57.65 (2.75) | **79.88** (2.65) | 0.76 (0.02) | 0.20 (0.01) | 0.16 |
|  | OpenL3 | 62.95 (1.42) | 61.64 (2.34) | 64.26 (2.84) | 0.69 (0.02) | 0.25 (0.01) | 0 |
|  | BYOL-A | 69.18 (1.36) | 61.75 (2.90) | 76.63 (2.19) | 0.76 (0.01) | 0.20 (0) | 0.13 |
| RF | Handcrafted | 66.41 (1.51) | 64.89 (3.39) | 67.92 (2.53) | 0.74 (0.02) | 0.21 (0) | 0.12 |
|  | eGeMAPS | 65.09 (2.04) | 61.53 (2.61) | 68.66 (1.76) | 0.72 (0.02) | 0.21 (0) | 0.13 |
|  | ComParE | 67.98 (2.18) | 64.58 (4.37) | 71.39 (2.22) | 0.73 (0.02) | 0.21 (0) | 0.14 |
|  | VGGish | 66.88 (2.03) | 64.89 (2.92) | 68.87 (2.36) | 0.73 (0.02) | 0.21 (0) | 0.13 |
|  | YAMNet | 67.98 (1.74) | 65.94 (3.02) | 70.02 (0.86) | 0.74 (0.02) | 0.21 (0) | 0.13 |
|  | OpenL3 | 68.08 (1.92) | 65.31 (3.22) | 70.86 (1.22) | 0.74 (0.02) | 0.24 (0.02) | 0.13 |
|  | BYOL-A | 67.35 (1.51) | 65.00 (3.82) | 69.71 (0.93) | 0.75 (0.01) | 0.21 (0) | 0.10 |
| XGBoost | Handcrafted | 65.04 (1.34) | 64.15 (2.58) | 65.94 (3.14) | 0.71 (0.01) | 0.27 (0.01) | 0.14 |
|  | eGeMAPS | 63.84 (2.63) | 62.68 (2.51) | 64.99 (4.39) | 0.70 (0.01) | 0.27 (0.01) | 0.12 |
|  | ComParE | 66.14 (1.37) | 62.79 (3.11) | 69.50 (1.92) | 0.70 (0.02) | 0.27 (0.02) | 0.16 |
|  | VGGish | 65.36 (2.08) | 64.47 (2.26) | 66.25 (3.08) | 0.70 (0.02) | 0.27 (0.02) | 0.17 |
|  | YAMNet | 65.31 (2.89) | 64.36 (3.22) | 66.25 (2.87) | 0.71 (0.02) | 0.27 (0.02) | 0.13 |
|  | OpenL3 | 64.21 (1.86) | 62.47 (2.24) | 65.94 (2.58) | 0.71 (0.02) | 0.27 (0.02) | 0.13 |
|  | BYOL-A | 66.04 (2.41) | 63.10 (2.39) | 68.97 (3.26) | 0.72 (0.02) | 0.26 (0.01) | 0.14 |
| MLP | Handcrafted | 67.92 (1.57) | 64.89 (2.78) | 70.96 (1.00) | 0.74 (0.02) | 0.21 (0.01) | 0.13 |
|  | eGeMAPS | 66.46 (2.75) | 61.33 (4.10) | 71.59 (2.18) | 0.73 (0.03) | 0.21 (0.01) | 0.12 |
|  | ComParE | 63.52 (2.96) | 59.66 (6.18) | 67.40 (5.17) | 0.68 (0.03) | 0.23 (0.01) | 0.09 |
|  | VGGish | 67.46 (2.69) | 63.74 (4.77) | 71.17 (0.91) | 0.74 (0.02) | 0.21 (0.01) | 0.14 |
|  | YAMNet | 68.97 (1.70) | 64.58 (4.61) | 73.37 (2.14) | 0.76 (0.01) | 0.20 (0.01) | 0.07 |
|  | OpenL3 | 63.58 (2.09) | 59.23 (5.56) | 67.93 (2.44) | 0.69 (0.02) | 0.23 (0.01) | 0.06 |
|  | BYOL-A | 68.45 (2.23) | 63.21 (3.21) | 73.70 (3.30) | 0.75 (0.02) | 0.20 (0.01) | 0.13 |

**Fig. 5:**
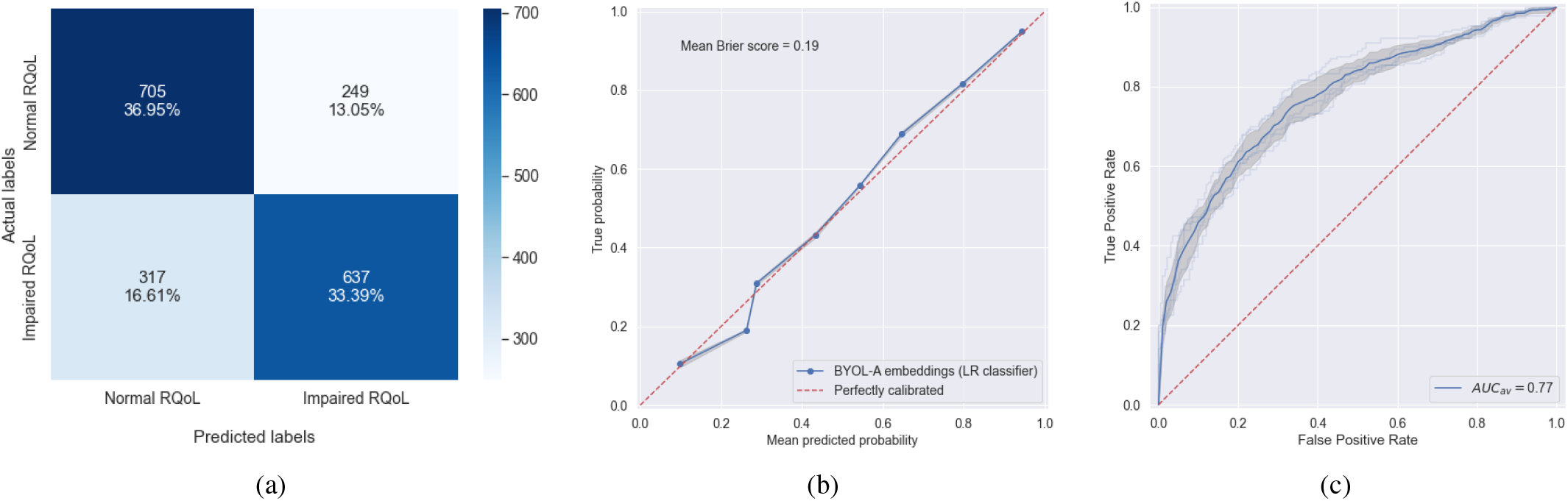
Performance of the best model (fused BYOL-A deep audio embeddings and socio-demographic/clinical features, trained with logistic regression classifier): a) Confusion matrix; b) Probability calibration curve; and c) ROC curve. Light blue lines denote the ROC curves across 5 cross-validation folds, whereas a thick blue line represents the average ROC curve. Standard deviation is highlighted with the shaded area.

Finally, since our objective was to quantify how much the vocal biomarkers increase the reliability of screening based only on subjective self-reports, NRI was used to estimate the improvement in performance after fusing vocal biomarkers with socio-demographic/clinical predictors. Table VI reveals that vocal biomarkers indeed improve the predictive capability of demographic and clinical variables for all acoustic features, with the biggest improvement measured by NRI of 0.19 for eGeMAPS features modeled with SVM.

## IV. Discussion

In this large international study, we have developed a digital voice-based biomarker for monitoring of RQoL using a combination of standard self-reported clinical information and voice-related features. We have shown that voice brings complementary information to improve the performances of the predictive model and increase the reliability of screening based only on subjective self-reports, reaching a full potential when both clinical and voice modalities are used conjointly in a multimodal setup.

RQoL has been evaluated from socio-demographic and clinical factors in various respiratory diseases, but mainly focusing on a single disorder, such as COPD [30], [31], asthma [32], idiopathic pulmonary fibrosis [33], or COVID-19 [34]. There were also attempts to investigate the effect of several respiratory diseases simultaneously on RQoL by using a multicase-control design, where the use cases were COPD, asthma, allergic and non-allergic rhinitis [35]. However, limited efforts were made to evaluate RQoL in the general population. A large five-year cohort study in Malawi was carried out to investigate the high prevalence of reduced lung function in Sub-Saharan Africa and its association with RQoL in the general population [36]. To establish a baseline for evaluation of RQoL from vocal biomarkers in the general population, we first estimated RQoL based on a number of socio-demographic (BMI, smoking habits) and clinical variables (day and night coughing, chest pain, sore throat, asthma, COPD). The feature importance analysis revealed that BMI is the most important socio-demographic variable. This confirms previous findings that BMI is significantly correlated with RQoL in COPD [37] and asthma [38], suggesting furthermore that RQoL of obese patients improves after weight reduction [37].

We further investigated whether digital biomarkers extracted from voice can act as a substitute for standard clinical measures estimated from questionnaires. Contrary to questionnaires which are mostly done during on-site clinical visits and can be tedious, voice recordings allow quick and easy-to-use data collection at patients’ homes; thus, substantially facilitating remote patient monitoring [39]. Our vocal biomarkers outperformed socio-demographic/clinical predictive factors by approximately 1.5% in terms of accuracy, confirming their potential to be a surrogate for clinical measures. The best-performing features are BYOL-A, which are general-purpose audio representations extracted with a model pretrained on a large amount of out-of-domain audio data in a self-supervised manner, i.e. requiring no annotations [28]. After freezing the convolutional layers, only the classification head is fine-tuned with the sustained vowel phonation collected within the Colive Voice study. This allows training the deep neural network models even with limited available voice data, and furthermore enables deploying for real-time inference, in applications that require low latency. However, deep audio embeddings such as BYOL-A suffer from limited interpretability, which might be an issue in a clinical application. Therefore, trade-off between performance and interpretability has to be considered when selecting the audio features.

Finally, fusing clinical and voice features in a multimodal setup allows focusing on different aspects of RQoL, localizing a broad range of information extracted from different modalities, and enabling more robust prediction models. The fusion of audio features with textual (word embeddings) and vision features (facial action units) has already been shown to improve the performance of unimodal approaches for the detection of clinical depression [40], [41]. A deep multimodal fusion model that learns indicators of Alzheimer’s disease from audio and text modalities, as well as disfluency features, increases the predictive power of audio features [42]. Fusion of speech, handwriting and gait data enables accurate evaluation of neurological state in different stages of Parkinson’s disease [43]. To the best of our knowledge, there were no previous attempts to combine voice features with clinical data for application in healthcare. By using intermediate feature level fusion we proved that voice features and clinical variables extracted from self-administered questionnaires are indeed complementary, leading to improved performance in comparison to both unimodal approaches by almost 5% in terms of accuracy, and up to 7% in terms of AUC. The intermediate fusion has an advantage in flexibility of extracting marginal representations appropriate for each modality, and arguably reflects more closely the relationships between the modalities [44]. To avoid producing high-dimensional joint feature representations, PCA was used to reduce the dimensionality of feature vectors coming from different modalities to the same length.

To further evaluate not only the ability of the model to accurately predict the class labels, but also the associated probability, the Brier score was used. The well-calibrated model is neither underconfident, nor overconfident, i.e. the true frequency of the positive label (impaired RQoL score in our case) against its predicted probability is approximately linear. This is confirmed by a solid average Brier score, and a calibration curve that does not deviate substantially from the perfectly calibrated model.

A major strength of this study is the fact that the dataset is acquired via a mobile app at participants’ homes, i.e. in uncontrolled conditions close to real-world circumstances. This confirms the feasibility of using a digital voice-based biomarker to provide quantitative measurements of RQoL, and enable regular remote monitoring in real life without relying on costly, invasive or cumbersome equipment; thus, facilitating personalized and more timely treatment, according to the patient’s needs and general health status.

However, a crowdsourced data collection poses multiple challenges and could be also observed as a limitation. There is a risk of acquiring low-quality answers from the self-administered questionnaires and introducing noise in the data, making it more difficult to infer the ground truth labels. We mitigated this risk by using a well-known, clinically validated questionnaire to assess RQoL. Recording voice using multiple devices, different qualities of microphones, and various recording conditions make data collection additionally challenging, resulting in different quality of audio recordings. For this purpose, we developed a proprietary data processing pipeline that harmonizes recordings and performs quality checks, but we cannot entirely exclude the possibility of having some low-quality recordings in our dataset.

## V. Conclusion

In this paper we developed a digital voice-based biomarker for monitoring RQoL in the general population. Our results confirm that vocal biomarkers can be a viable surrogate for standard clinical measures estimated from questionnaires, but the ultimate capacity is unlocked in a multimodal setup when clinical and voice data are used together. The best performance was obtained with a feature-level fusion of BYOL-A deep audio embeddings and socio-demographic/clinical variables, reaching an accuracy of over 70% and AUC of 0.77, a performance boost of 5% in comparison to acoustic features extracted from voice only.

The proposed approach facilitates rapid screening and represents a step towards the development of scalable, non-invasive, easy-to-use and low-cost solutions for remote monitoring of respiratory health status.

## Data Availability

All data produced in the present study are available upon reasonable request to the authors.

## Acknowledgment

Colive Voice study is funded by the Luxembourg Institute of Health. The funder played no role in the study design, data collection, analysis and interpretation of data, or the writing of this manuscript.

We would like to thank all participants that contributed to Colive Voice study, as well as our partners for their help in recruiting new participants. Special thanks go to Aurélie Fischer, Philippe Kayser, Luigi De Giovanni, Michael Schnell and Aurore Dobosz for their substantial contribution to the Colive Voice study.

## Footnotes

1 https://www.colivevoice.org)

2 https://app.colivevoice.org/

